# Estimating the proportion of coronavirus disease 2019 (COVID-19) cases among households in France: a cross-sectional study based on individuals with myocardial infarction history

**DOI:** 10.1101/2020.06.19.20135418

**Authors:** Laurie Fraticelli, Julie Freyssenge, Clément Claustre, Mikaël Martinez, Abdesslam Redjaline, Patrice Serre, Thomas Bochaton, Carlos El Khoury, on behalf of the RESCUe-RESUVal team

**Affiliations:** RESCUe & RESUVal Network, Lucien Hussel Hospital, Vienne, France; Laboratory Systemic Health Care, EA4129, University of Lyon, Lyon, France; University Claude Bernard Lyon 1, HESPER EA 7425, Lyon, France; Emergency Department, Hospital Center du Forez, Montbrison, France; Emergency network Loire Ardèche Nord (REULIAN), Hospital Center Le Corbusier, Firminy, France; Emergency Department, Hospital Center Firminy, Firminy, France; Emergency Department, Hospital Center Fleyriat, Bourg-en-Bresse, France; Emergency and Critical Cardiac Care, Cardiologic hospital, Hospices Civils de Lyon, Bron, France; Medipole Hopital Mutualiste, Emergency Department and Clinical Research Unit, Villeurbanne, France

**Author notes:** Corresponding author (LF). Membership of the RESCUe-RESUVal team is provided in the Acknowledgments.

## Abstract

**Background:** Previous studies have identified that adults with cardiovascular diseases were disproportionately associated with a significantly increased risk of a severe form of COVID-19 and all-cause mortality. We aimed to describe the diagnosed COVID-19 cases and to estimate the symptomatic and asymptomatic suspected cases among individuals with pre-existing myocardial infarction myocardial infarction and their relatives in lockdown period. Methods: We conducted a two-week cross-sectional telephone survey, from May 4 to May 15, 2020, including all households with at least one individual with pre-existing cardiovascular disease in the past two years. We defined a suspected COVID-19 case when living with at least one individual tested positive to COVID-19, or when an individual has been in contact with a suspected or confirmed case since the March 1rst, or when a relative from the same house has been hospitalized or deceased for COVID-19.

**Results:** We observed high rates of compliance with health measures during the lockdown period, regardless of age or risk factors. Among individuals with myocardial infarction history, two were COVID-19 confirmed, 13·37% were suspected (94/703) of whom 70·21% (66/94) asymptomatic.

**Conclusions:** Individuals with myocardial infarction history presented different symptoms association with more respiratory signs. This population, which is older and associated with more comorbidities, is exposed to a high risk of complication in the event of contamination. Infection rates are relevant to adjusting the management of emergency departments in our region.

## Introduction

With more than four million confirmed cases worldwide, the novel coronavirus disease 2019 (COVID-19) has caused more than 350,000 deaths since December 2019. When European countries are closed to unlock-down populations, the need for early and accurate diagnosis for suspected cases become obvious for effective management and for keeping control of the disease spreading [1].

Virological tests (reverse transcription polymerase chain reaction, RT-PCR) have routinely been used to confirm diagnosis for suspected cases, providing results within a few hours. But recent studies highlighted a high false-negative rate, with a sensitivity as low as 38%, which is not better than chance [2]. Although serological tests can inform if individuals were exposed to the virus and if they presumably developed immunity, the poor analytical performance can create confusion and may lead to false reassurances, especially when carried out on large populations that have yet to be exposed to the virus and in the absence of a gold standard comparative method [3]. When COVID-19 diagnosis is confirmed, chest computed tomography (CT) scan has a central place in the management of respiratory symptoms but cannot be generalized at the scale of the general population. We are therefore faced with the need to find a more reliable method for estimating prevalence.

We may have underestimated the potential of population surveys in identifying suspected COVID-19 cases. Existing surveys were designed to assess qualitative data such as risk perception, social isolation or behavioural disorders. Since the symptoms are now well documented in the literature [4], we hypothesized that surveys could also be useful to estimate the suspected and asymptomatic COVID-19 cases. Moreover, the most severe forms of COVID-19 and overall risk of all-cause mortality were disproportionately associated with older adults because age and pre-existing conditions [5–8].

We aimed to describe the diagnosed COVID-19 cases and to estimate the suspected cases among individuals with pre-existing myocardial infarction, selected from an observational registry, as well as symptomatic and asymptomatic cases among their relatives during the lockdown period. We secondary aimed to describe risk perceptions and precautionary behaviors.

## Materials and Methods

### Study design

We conducted a two-week cross-sectional telephone survey, from May 4 to 15, 2020, corresponding to the last week of lockdown in France and the five following days (the average period of incubation [9]). The sample comprised households with at least one individual with pre-existing myocardial infarction occurring in the Auvergne Rhône-Alpes region in France.

### Sample selection

The eligible households were identified through inclusions in the OSCAR registry, a multicentric prospective observational registry (for “Observatoire des Syndromes Coronaires Aigus dans RESCUe”) of the regional emergency cardiovascular network (RESCUe) [10]. Funded by the Regional Agency for Health (Agence Régionale de Santé Auvergne Rhône-Alpes), the network covers 3 million inhabitants located in the second most important region in France, including 10 large-volume hospitals, representing more than 400 percutaneous coronary interventions per year. The OSCAR registry was approved by the French National Commission of Informatics and Liberties (Commission Nationale de l’Informatique et des Libertés [CNIL]) (number 2013090 v0) and all the participants gave informed and oral consent. Patients included were associated with persistent chest pain with ST segment elevation of at least a two millimeters in at least two continuous leads. We extracted the 1,164 myocardial infarctions listed in the OSCAR registry, which occurred between September 3, 2018 and December 10, 2019, sent home and successfully reached by telephone at least once for cardiological follow-up. With two years of hindsight and the inclusion criteria chosen, we have increased the chances of successfully reaching households by telephone. We excluded the patients discharged from dependent elderly homes and the household where the patient was deceased in the meantime.

### Investigator training

A total of 17 investigators were involved in the telephone interviews. They were trained by an experienced telephone operator for two two-hours meetings. The training sessions included information on the context to setting up the survey, the methodology, the construction of the sampling frame, and eligibility criteria, the conduct of the questionnaire and the contact phase. After a brief presentation of the study, investigators collected the oral consent from the first respondent to allow the collection of anonymous data for all individuals living in the household during the lockdown period. In accordance with French regulations, an information note was addressed by email or postal mail to the household to explain the purpose of the study and the rights of after data collection. Interviews took place between 10 am and 12 pm, and between 1 pm and 6 pm. If no response was received, three other telephone attempts were made at different time slots and days.

### Data collection

The survey was based on a scoping review of the PubMed scientific literature. The investigator collected information from the first respondent. The questionnaire comprised a common part relating to the household in general, and another part relating to each individual living in the household during the lockdown period. The common part relating to the household consisted in identifying the place of residence (zip code and name), the number of individuals and any possible regular contacts of a third person (home nurse or home helper). In addition to the items on COVID-19 symptoms observed since March 1, the questionnaire for individuals comprised sociodemographic items (age, sex, weight, height, occupation), the respect of precautionary behaviours (physical distancing, contact outside home, number of outings per week), pre-existing comorbidities and treatments, travels to high-risk areas in France or abroad, and the results of nasal or blood testing or chest CT scan. Moreover, it included the delivered, reported or renounced consultation during the lockdown period, and if another individual of the household was hospitalized or deceased for COVID-19.

### Definitions and assumptions

We defined a suspected COVID-19 case when the individual was living with at least one individual who tested positive to COVID-19 [11], or when had been in contact with a suspected or confirmed case since March 1, or when a relative from the same household, not present at the time of the survey, was hospitalized or deceased of COVID-19. We assumed that when an individual is confirmed or suspected to be COVID-19 positive, the whole household is considered to be suspected of being infected with COVID-19.

### Statistical and geographical analysis

To determine the representativeness of the study sample, we used the open source data from the French Institute of Statistics and Economic Studies (INSEE) to compare the respondents included in the survey to the inhabitants living in the same area, based on a two-stage approach by age [5] and sex [12].

We provided baseline characteristics in numbers and percentages for categorical variables and medians and interquartile ranges for continuous variables. Bivariate analyses were assessed using the Pearson Khi^2^ test for categorical variables and the non-parametric Wilcoxon rank test for continuous variables. Statistical analyses were performed using R 3.6.2 software. The level of significance was set at a p value <0·05.

The missing data represented less than 1%, except for the body mass index (10·12%). We specified the denominator in Table 1 when different from the total number.

**Table 1:**
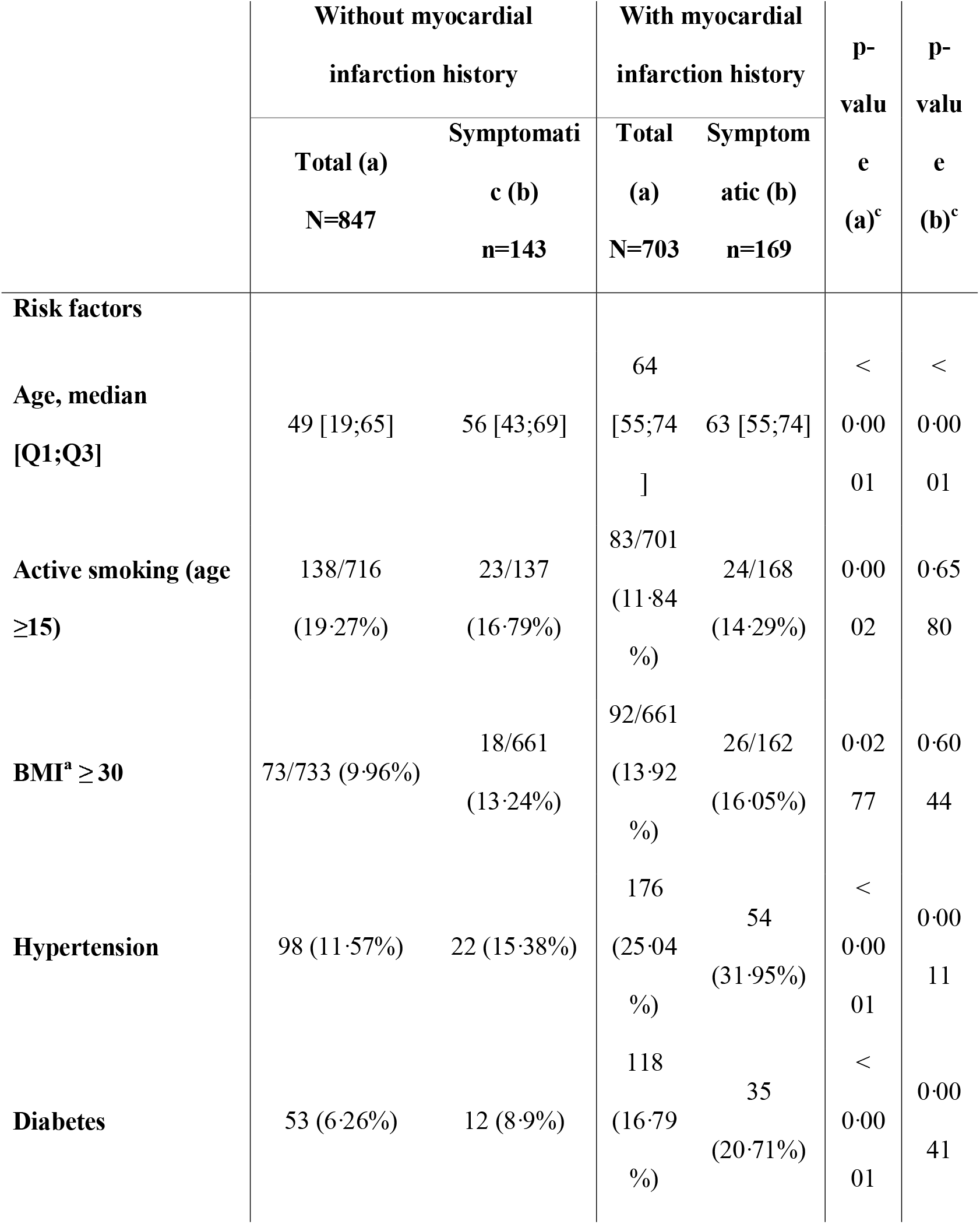
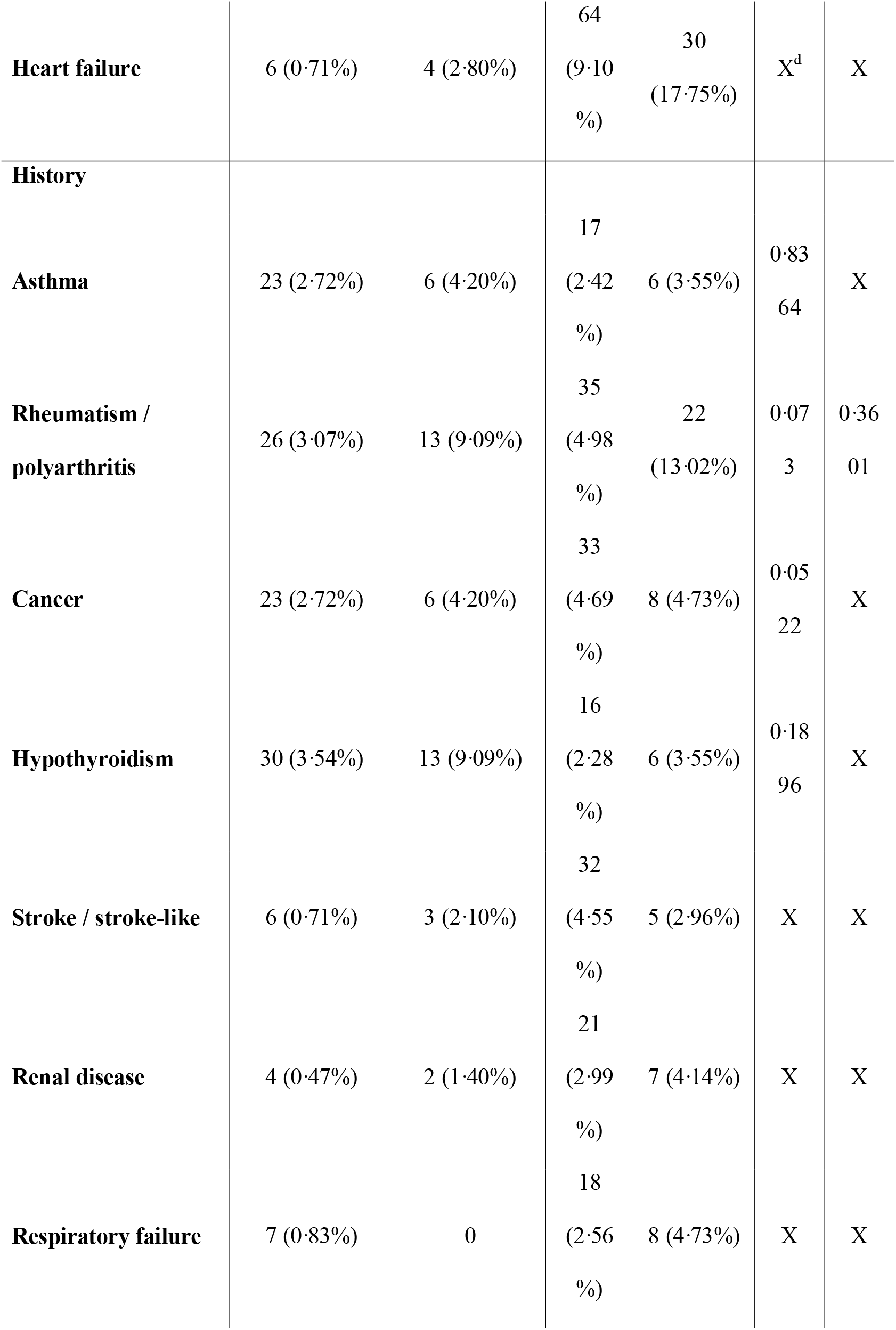
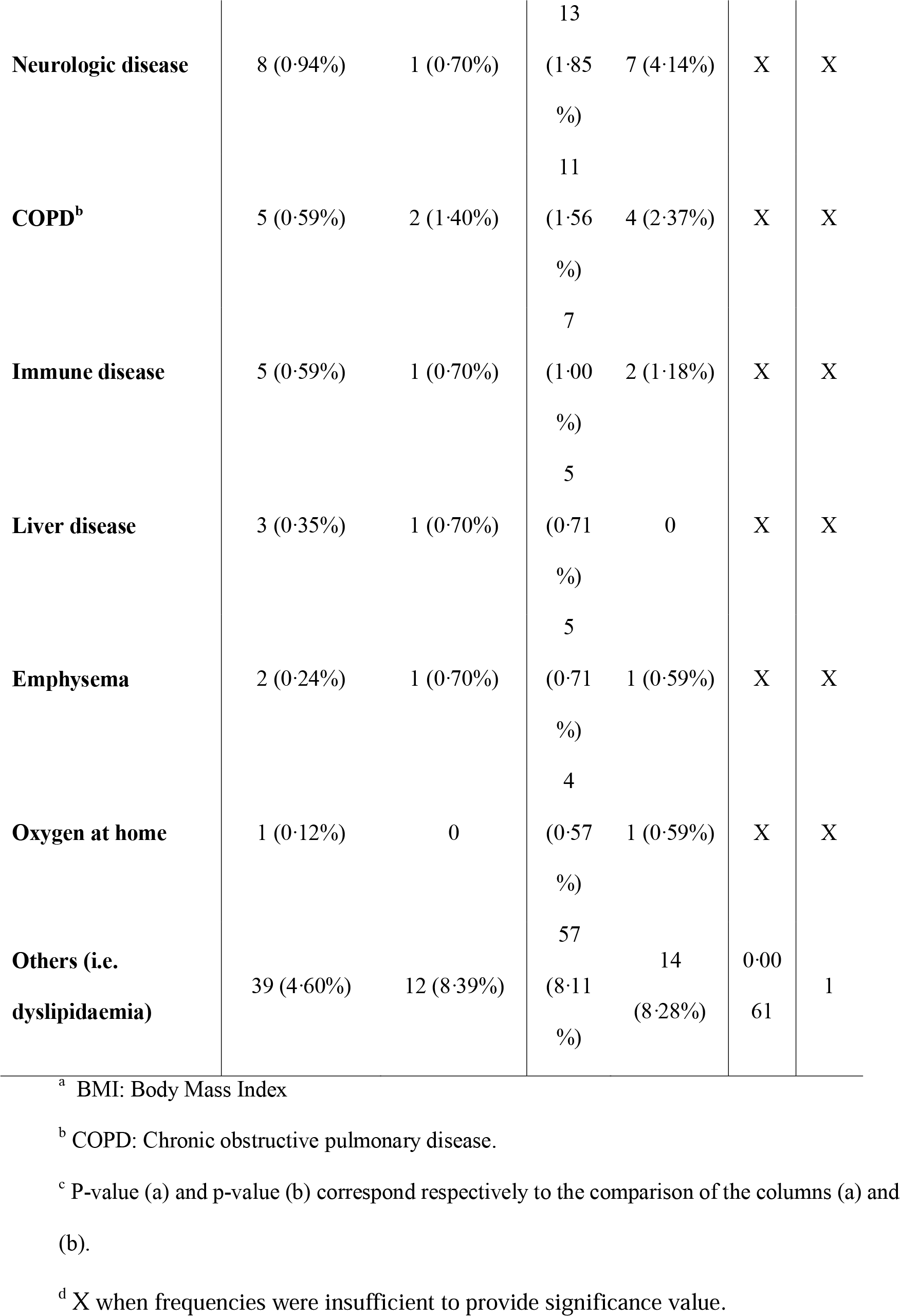
Risk factors and history of the n=1,552 individuals (two missing values).

We focused on symptom associations in order to understand the links between clinical signs among suspected COVID-19 cases. We used a network approach [13] where the nodes represented an association of symptoms (reported by at least two individuals), linked by the same shared symptom(s). We compared individuals with and without myocardial infarction history.

## Results

### Inclusions statement

We identified 1,164 eligible households from the OSCAR registry. The investigators made 1,052 call attempts in a two-week telephone survey, with an average of 1·63 average calls per household. A total of 668 households gave their consent to participate, representing 1,552 individuals (Fig 1). The participation rate was about 88·70% (668/753). We observed 134 individuals living alone during the lockdown period (i.e. 20·06% of the households). We also reported 703 individuals with myocardial infarction history (i.e. 45·30% of the individuals). The proportions of women (50·06%) and men (49·94%) were balanced. However, the study sample included older individuals compared to the resident households of the area (Fig 2). To illustrate, we emphasized that the sample was composed of three times less of 30-44 years old men compared to the inhabitants of the survey area and twice as many men between 60-64 years old.

**Fig 1:**
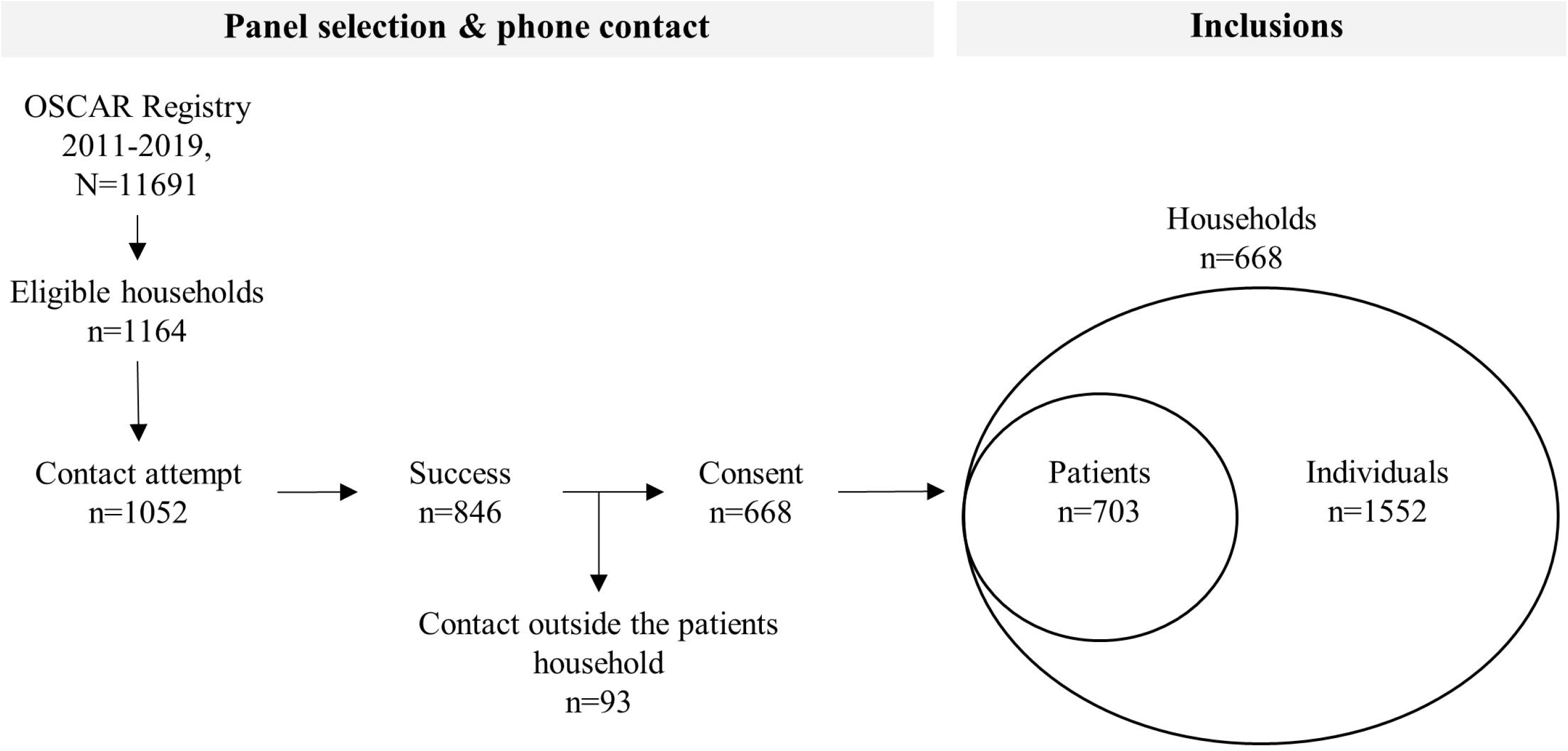
Flowchart of the sample selection and inclusions.

**Fig 2:**
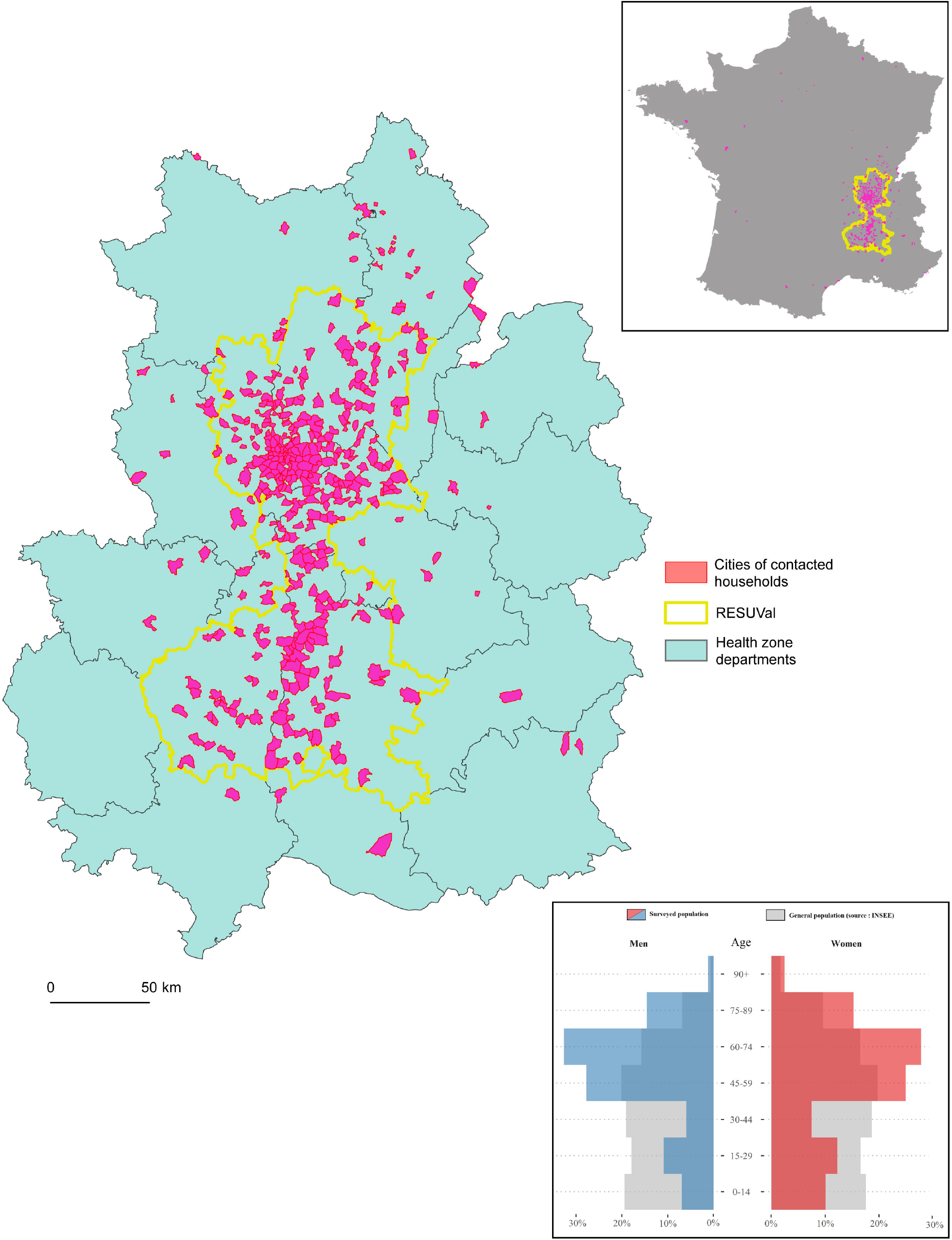
Territorial coverage and age and sex representativeness of the households included in the telephone survey (N=1’552 in France for n=1’507 in the RESUVal area and its adjacent departments).

### Risk behaviour and risk factors

We observed high rates of compliance with health measures, regardless of age or risk factors. Among the 1,373 individuals who did not work at their usual workplace (i.e. 88·47%), 98·69% were confined, 98·69% maintained physical distances, 29·42% had no contact with individuals outside their household during the lockdown period and 23·89% went out only once per week. Only 14·21% of the households were regularly visited by a caregiver or a nurse. Among the individuals at risk (age ≥ 60 years old with at least one comorbidity) (n=709), we observed 98·59% of compliance with lockdown and 99·15% with physical distances.

Less than a third of individuals was vaccinated against influenza A H1N1 (32·86%). This proportion reached 49·93% among individuals with myocardial infarction history (recommended by the 2017 ESC guidelines [14]). Nearly a quarter of the sample presented at least one COVID-19 symptom (20·62%). We observed more symptomatic individuals with myocardial infarction history (24·47% vs 17·24%, p=0·0006). These groups of individuals were significantly associated with different risk factors and comorbidities in Table 1.

### Symptomatology associated with COVID-19

We observed that 20·23% of individuals were symptomatic, of whom 49·38% reported only one symptom. The most frequent symptoms were cough (29·62%), headaches (27·07%), runny nose (25·16%), unusual fatigue (20·70%), fever (18·47%), and sore throat (16·61%). We identified 37 associations reported by at least two individuals, 20 associations for individuals with myocardial infarction history (n=95) and 17 without myocardial infarction history (n=97) (Fig 3). Individuals with myocardial infarction history were more associated with isolated symptom compared to other individuals, accounting for 40% (38/95); headache (n=12), chest pain (n=8), muscle pain (n=5) abdominal pain (n=5), and dermatological lesions (n=2). To illustrate, in individual without pre-existing myocardial infarction, headaches (n=31) were frequently associated with other symptoms like runny nose (n=7) cough (n=12), sore throat (n=3), fever (n=2), unusual fatigue (n=3), muscle pain (n=2). In individuals with myocardial infarction history, the most associated symptom was cough (n=31) with sore throat (n=5), runny nose (n=5), unusual fatigue (n=2), and fever (n=4).

**Fig 3:**
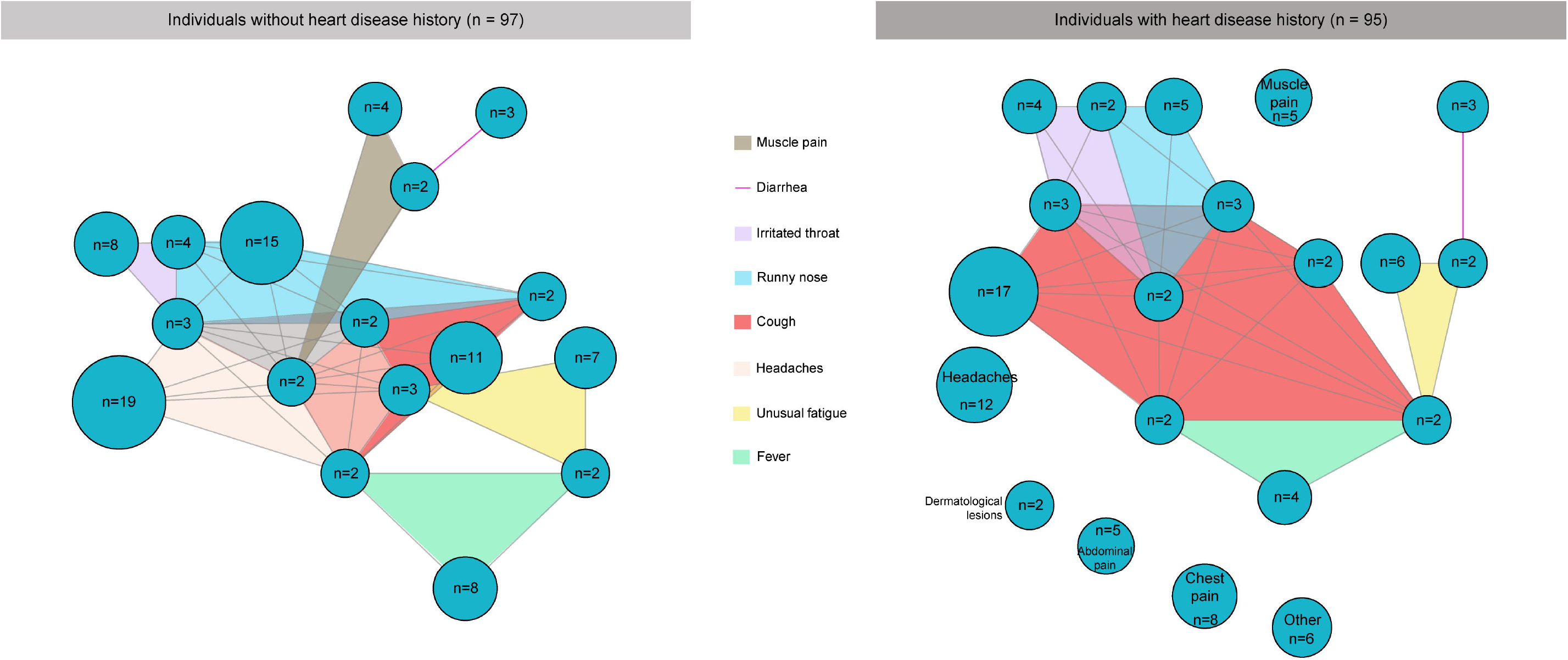
Symptom association network: comparison of clinical signs between individuals with and without heart disease history.

### Consultations during lockdown period and diagnostic tests

Only 38·85% of the symptomatic individuals consulted a general practitioner because of their symptoms; 68·03% of them went to the doctor’s office, 15·57% used a telemedicine service or 16·39% visited the emergency department.

Nearly a quarter of individuals (23·78%) had their appointments rescheduled at physician’s initiative, but these delays were more common among individuals with myocardial infarction history (36·84% vs 12·99%, p<0·0001). We also noted that 7·54% of them decided on their own to cancel an appointment (vs 3·78%, p=0·0018), and 9·10% to report it (vs 2·72%, p<0·0001). In 76·07% of those cases, the appointment was related to medical follow-up (vs 67·27%, p=0·3026).

Only 43 individuals (2·77%) were tested. The test processes were RT-PCR in 55·81%, chest CT in 23·26% and blood tests in 44·19%. Only 34 tests were performed among the 314 symptomatic individuals (10·83%) with four positive results leading to one hospitalisation (Fig 4) (one by RT-PCR, one by blood tests, one by chest CT scan and one by the three tests). We observed that 0·26% of individuals were confirmed COVID-19 cases (4/1,552) and estimated that about 15·59% were suspected cases (242/1,809) of whom 74·79% were asymptomatic cases (181/242). In the subgroup of individuals with myocardial infarction history, two individuals were COVID-19 confirmed among four positive tests, 13·37% suspected cases (94/703) of which 70·21% (66/94) were asymptomatic suspected cases.

**Fig 4:**
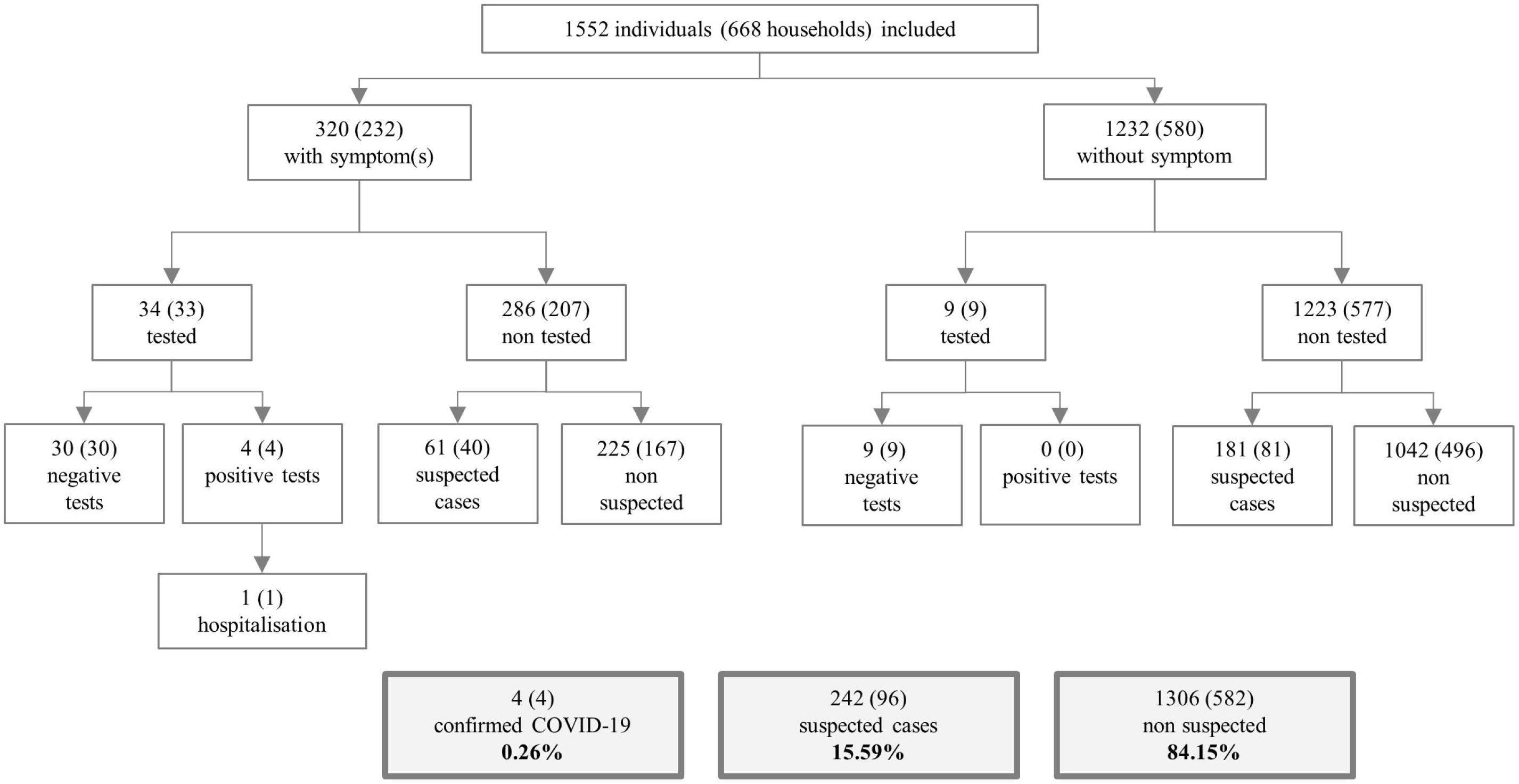
Flowchart of testing process among individuals and households sampled.

## Discussion

Since the prevalence of COVID-19 is unknown, we considered the cross-sectional study to be a useful means of understanding the pandemic situation at a given time and place [15]. While time series models allow us to understanding the trends of the outbreak and to estimate the different epidemiological stages, we still need a reliable and turnkey solution to evaluate the situation in a high-risk population. Conducted at the end of the lockdown period in France, we presumed that this survey is relevant over a limited area due to the uneven distribution of infection rates across France. Indeed, the Auvergne Rhône-Alpes region was moderately affected compared to the Grand Est region [16]. Nevertheless, outcomes such as risk behaviours and symptoms remain declarative responses and subject to approximations.

Although the sample was not representative of the housing estate, we included an exhaustive population of myocardial infarction history, focusing on middle-aged and elderly individuals. Literature has established that the latter are associated with higher mortality compared to young or middle-aged individuals [17]. The sample study would be associated with higher risk of complications or hospitalization if it included 45·30% of individuals with myocardial infarction history. Previous studies stated that 16·40% of the hospitalized COVID-19 patients were associated with cardio-cerebrovascular diseases [18]. In addition to these risk factors, we observed in our sample that included individuals were active smokers in 15·60%, obese in 11·84% and diabetics in 11·03% [19–21].

The high participation rate of approximately 88·70% (668/753) shows that telephone survey is a study with a suitable and feasible design to address such research question in a short period of time [22]. This proportion is probably due to the population availability during the lockdown period and to their high interest in this unprecedented pandemic situation. If we had sent out a paper questionnaire by postal envelope or surveyed through an online questionnaire, the participation rate would have been much lower [23]. We also considered that the period of the survey was appropriate – the last week of lockdown and the first week post-lockdown - because we are taking a step back on the incubation period of COVID-19 before the lockdown and on the precautionary behaviours during the lockdown period.

The study sample also provided estimations on adherence to precautionary measures during the lockdown period with a high compliance rate in our high-risk population. This compliance with precautionary measures certainly explains the only one hospitalization and the absence of deaths. We assume that relatives were susceptible to adapting their behaviour and therefore not putting the individual with myocardial infarction history at risk of infection. Nevertheless, we cannot assume that precautions are observed in the whole population, especially among healthy individuals.

We provided a network approach to understand the different clinical signs associated with suspected cases of COVID-19. We observed that individuals with myocardial infarction history were most likely to have respiratory symptoms such as cough, sore throat, runny nose, and fever, whereas other individuals presented non-specific signs (i.e. headaches, muscle pain).

## Conclusions

We conducted a cross-sectional telephone survey by selecting households from a prospective observational registry of individuals with myocardial infarction history. We observed a low proportion of symptomatic individuals tested with 10·62% (34/320). We estimated that approximately 13·37% (94/703) of the cases were suspected, of which 70·21% (66/94) were asymptomatic. A telephone survey is a relevant tool for assessing the number of suspected cases on a limited area, based on the presence of associated symptoms and confirmed or suspected infected contacts. These estimates advance our knowledge to better prepare for future crises.

## Data Availability

Data are available on demand.

## Acknowledgements

The RESCUe Network is funded by the Regional Agency for Health from Auvergne-Rhône-Alpes region (Agence Régionale de Santé Auvergne-Rhône-Alpes). The team RESCUe-RESUVal has participated to the data collection; Sylvie Besnier, Magali Bischoff, Aurélie Couzon, Nicolas Eydoux, Marie-Anne Gaud, Violaine Larat, Mylène Masson, Marie Mirnik, Julie Morel, Conception Nunez, Alexandra Peiretti, Catherine Vincent. The co-authors thank Emeline Moderni for the proofreading of English.

